# Bimanual upper limb task practice and Transcutaneous electrical stimulation enhance spinal plasticity and hand function after chronic cervical spinal cord injury

**DOI:** 10.1101/2025.01.06.24319679

**Authors:** Antonio Capozio, Samma T Chowdhury, Samit Chakrabarty, Ioannis Delis, Maria Horne, Manoj Sivan, Parag Gad, Raymond Holt, Ronaldo Ichiyama, Sarah L Astill

**Affiliations:** School of Biomedical Sciences, University of Leeds, Woodhouse Lane, Leeds LS2 9JT, United Kingdom; Department of Psychology, Edge Hill University, Law and Psychology building, St Helens Rd, Ormskirk L39 4QP, United Kingdom; Faculty of Medicine and Health, School of Healthcare, University of Leeds, Woodhouse Lane, Leeds, LS2 9JT, United Kingdom; Leeds Institute of Rheumatic and Musculoskeletal Medicine, Woodhouse Lane, Leeds, LS2 9JT, United Kingdom; SpineX Inc., CA, 90064, Los Angeles, United States of America; School of Mechanical Engineering, University of Leeds, Woodhouse Lane, Leeds, LS2 9JT, United Kingdom

**Keywords:** spinal cord injury, spinal stimulation, task practice, hand function, neural plasticity

## Abstract

Injuries to the spinal cord at the cervical level can lead to loss of upper limb function. Recent work suggests that combining functional task practice with Transcutaneous electrical stimulation of the spinal cord (TCES) can increase strength and upper limb function in people living with chronic cervical spinal cord injury (cSCI). Participants (*n* = 5) were randomly assigned to: Group_1 (*n* = 3) receiving one month of upper limb task practice (ULTP) followed by one month of upper limb task practice paired with spinal stimulation (ULTP+TCES); Group_2 (*n* = 2) receiving one month of ULTP+TCES followed by one month of ULTP. Changes in hand function (assessed via the Graded Redefined Assessment of Strength, Sensibility), independence and quality of life were investigated after each intervention and at three-months follow-up. In addition, we assessed cortical (via Transcranial Magnetic Stimulation) and spinal (via single-pulse TCES) excitability at those same time points. For Group_1: improvements in hand function from baseline were observed after ULTP+TCES (*p<*0.001) and at follow-up (*p=*0.017); quality of life increased between baseline and after ULTP (*p=*0.002), ULTP+TCES (*p<*0.001) and at follow-up (*p=*0.013); spinal excitability increased from baseline to after ULTP+TCES (*p*<0.001). For Group_2: improvements in hand function from baseline were observed after ULTP+TCES (*p<*0.001), ULTP (*p<*0.001) and at follow-up (*p<*0.001); corticospinal excitability increased from baseline to after ULTP (*p*=0.013); spinal excitability increased from baseline to after ULTP+TCES (*p*<0.001) and the increase persisted 3 months later at follow-up (*p*=0.04). Our findings demonstrate that non-invasive spinal stimulation paired with task practice can improve hand function more than task practice alone in people living with a cSCI. In addition, we suggest that spinal plasticity induced by spinal stimulation is a potential neural substrate for the attained improvements in hand function.

## Introduction

Spinal cord injury (SCI) can lead to devastating effects on motor function, independence and quality of live. Injuries to the cervical levels of the spinal cord are more common than injury to the lower segments of the spinal cord, thus tetraplegia is more common than paraplegia[1]. Tetraplegics rank regaining arm and hand function as their main priority for rehabilitation, five times greater than bowel, bladder, sexual or lower extremity function[2]. Thus, identifying and optimising therapies to restore functional arm and hand recovery is an important clinical, economic and social goal[3]. Task-specific training is currently the most effective evidence-based way of augmenting upper limb rehabilitation after SCI, especially when completed concurrently with a neuromodulatory technique inducing neural plasticity[4, 5]. One such therapy which has shown promising results is transcutaneous electrical stimulation (TCES) of the spinal cord[6]. In people living with cervical SCI, TCES paired with intensive functional training promoted recovery of upper limb strength and function as measured via the Graded Redefined Assessment of Strength, Sensibility, and Prehension (GRASSP) test[7]. In the study by Inanici et al.[7], following an initial month of training, participants completed a month of training paired with stimulation followed by one further month of training alone and one of training paired with stimulation. Similarly, Moritz et al., demonstrated that two months of stimulation paired with task practice following two months of rehabilitation alone improve upper limb function in the majority of participants[8]. Despite these promising results, no direct comparison between task practice and task practice with spinal stimulation has been presented so far and no evidence that spinal stimulation can improve upper limb function even without a prior period of task practice, a finding which could improve the design of rehabilitation paradigms. In addition, while studies reported increases in function as evidenced by changes in GRASSP scores or similar test batteries, transfer of improvements to daily activities such as grasping and lifting objects is yet to be investigated.

While the evidence for a positive effect of TCES on strength and function is ubiquitous, there is limited knowledge of the neural substrates underpinning the behavioural changes[9]. Changes in neural excitability occurring at the cortical and spinal level after cervical TCES seem to occur at both the acute (e.g. within-session) and longer (e.g. after multiple sessions) timescales[10]. At the acute level, TCES increases corticospinal excitability as measured via Transcranial Magnetic Stimulation (TMS) in healthy participants and people living with SCI[11], but only in the absence of a high-frequency carrier component[11, 12]. Spinal excitability, assessed via transcutaneous single pulses of spinal stimulation at the cervical level, was also found to be increased: after one session[11] of TCES alone; after 4 weeks[13] and 8 weeks[14] of spinal combined with task-specific hand training. To our knowledge, no study up to date have investigated cortical and spinal changes in excitability occurring throughout a rehabilitation protocol which alternates task practice and task practice with stimulation. Moreover, we are not aware of any study reporting the neurophysiological effects of TCES over a longer timescale at the end of the protocol.

Given the above gaps in the literature, the objectives of the current study were: (1) to compare the effects of one month of ULTP followed by one month of ULTP+TCES with the effects of one month of ULTP+TCES followed by one month of ULTP on hand function, independence and quality of life; (2) to assess changes in corticospinal excitability occurring after each intervention and at three months after the end of the interventions; (3) to assess changes in spinal excitability occurring throughout the intervention and at three/months after the end of the intervention; (4) to determine if the observed effects on hand function are paralleled by changes in force production while participants complete grasping and lifting tasks.

## Materials and methods

### Participants

Five participants (*M* ± *SD* = 48 ± 10 years; females = 5) living with a cervical spinal cord injury volunteered for the study. Participants characteristics are described in Table 1. Inclusion and exclusion criteria are listed in Table 1. Group allocation was randomised and counterbalanced across participants. All participants gave written informed consent to experimental procedures approved by the HRA and Health and Care Research Wales (HCRW) (REC reference: 22/NW/0171) and conformed to the Declaration of Helsinki. The study was registered with ClinicalTrials.gov (NCT05801536).

**Table 1.** Demographics characteristics of the participants. AIS = American Spinal Injury Impairment Scale. ULTP = Upper Limb Task Practice, GROUP = 1, ULTP first; 2, ULTP+TCES first.

| Participant | Sex | Age (years) | Injury level | Time since injury (years) | AIS | GROUP |
| --- | --- | --- | --- | --- | --- | --- |
| 242 | F | 51-55 | C6-C7 | 8 | C | 1 |
| 192 | F | 31-35 | C5-C6 | 16 | C | 2 |
| 650 | F | 61-65 | C3-C8 | 6 | D | 1 |
| 618 | F | 41-45 | C5-C6 | 2 | C | 2 |
| 528 | F | 51-55 | C4-C6 | 30 | D | 1 |

### Study design

All participants attended the lab for a total of 34 sessions over the course of 25 weeks. On the first 2 weeks, participants completed two sessions per week (sessions 1 to 4): on each of these days they completed either a Functional assessment or a Neural assessment (Figure 1). We chose to include two baseline sessions to ensure stability of our primary baseline measure (e.g., GRASSP) prior to the intervention (*t(*9)=0.00 *p*=1). Participants were then randomly allocated to either Group_1, receiving ULTP first and then ULTP+TCES, or Group_2 receiving ULTP+TCES first and then ULTP in a crossover fashion according to a predefined randomisation list generated using a randomiser tool (GraphPad Prism; GraphPad Software Inc., CA, USA). Each of the interventions comprised of 4 weeks in which participants attended the lab for three sessions per week separated by at least 24 hours. At the end of the two intervention stages, participants completed a further two (one session of functional assessment, one session of neural assessment). Finally, participants were asked to attend the lab twice again (one session of functional assessment, one session of neural assessment) 3 months after their last intervention session to assess any prolonged effects of the two interventions outlasting the interventions.

**Figure 1.**
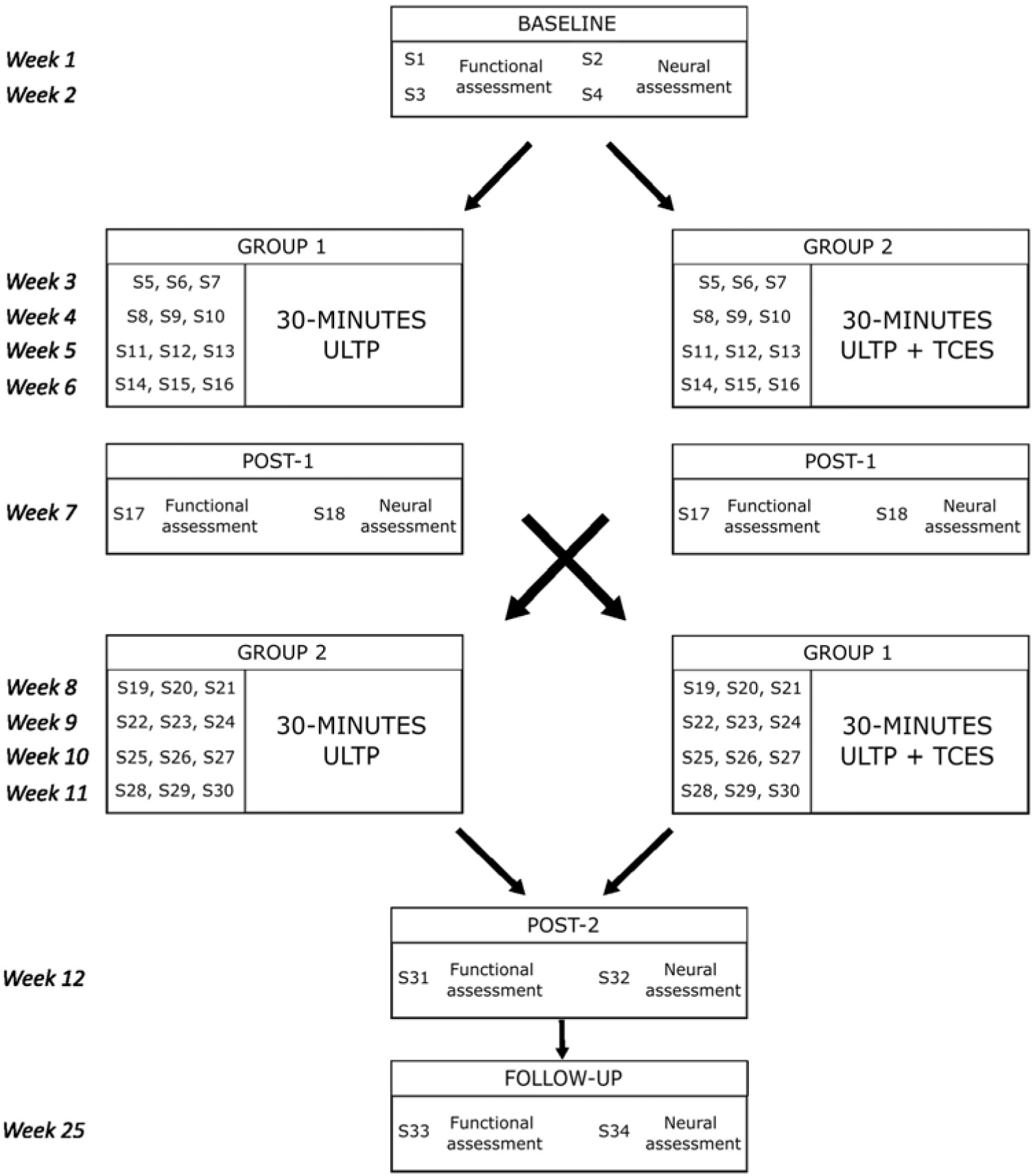
Experimental design. A randomised crossover study with outcome measures collected twice at baseline (Session 1-4), once after the first intervention (Post-1), once after the second intervention (Post-2) and once at three-months follow-up.

### Interventions

#### Upper limb task practice (ULTP)

The ULTP was designed by a clinician and a physiotherapist and comprised practice of unimanual and bimanual tasks that emphasize six categories of activities thought to represent different aspects of functional hand use (symmetrical, wherein both hands perform similarly and asymmetrical, wherein one hand performs the fine motor activity while the opposite hand stabilizes the object being manipulated, tasks)[5, 15]. The chosen activities were: independent finger movement, precision grip (pinch), pinch with object manipulation, power grip (grasp) complex power grip (involving object manipulation), finger isolation, and whole arm movement (Supplementary Fig. 1, *A*). Activities were scored by the researchers and task difficulty could be incremented/decremented over the following sessions according to participants’ performance. Completion of the ULTP was supervised by members of the experimental team, feedback was given by the researchers to discourage (or decrease) use of compensatory strategies such as tenodesis grasp. Rest periods were provided between each activity category if the participants required. The total time spent training for each session was approximately 30 minutes (five minutes for category), excluding potential rests.

### ULTP + TCES

#### Mapping procedure

During the first two neural assessment sessions, a mapping procedure was completed to establish the intensity and loci of stimulation for continuous TCES. First, the spinal segments to be stimulated were chosen according to the extent of the spinal lesion, with one cathodal electrode placed on the vertebral segment immediately above the lesion site and one immediately below it, between spinous processes; second, participants performed three maximal grip tasks with each hand, and the weakest (e.g. most affected) arm was identified as the one producing lower force values; third, in order to target preferentially the most affected arm, three loci of stimulation (midline, left to the midline, right to the midline) were tested on the same spinal segment for each cathode (Supplementary Fig. 1, *B*). Fourth, stimulus intensity was incremented until a clear increase in maximal force was observed in the most affected hand. Importantly, immediate and substantial increases in grip force were observed in all participants and mapping sessions; fifth, the locus for which the highest increase in force was observed was chosen for delivering TCES. At the beginning of each experimental session, we confirmed that stimulation at the chosen intensity produced increases in maximal grip strength in the most affected hand before the participants started to engage with the ULTP tasks and adjusted stimulation intensity if needed. Throughout the mapping procedure and while ramping up the stimulation intensity at the beginning of each intervention session, participants were asked to rate their perceived pain level from 0 to 10 using a visual analogue scale for pain, with 0 defined as “no discomfort at all” and 10 as “unbearable pain”. Stimulation was halted immediately if discomfort reached level 8 out of 10[12].

#### Use within the intervention

During the ULTP+TCES intervention sessions, TCES was delivered on continuous mode while participants completed the ULTP activities. TCES was delivered using the <u>TESCoN</u> device (SpineX Inc., Los Angeles, CA) using delayed biphasic blocks of pulses at a frequency of 30 Hz: each block contained 10 pulses of 100µs length (modulating frequency of 10 kHz). Stimulation was delivered via two 2.5 cm electrodes (Axelgaard, ValuTrode Cloth) as cathodes and two 5 × 10 cm electrodes over the iliac crests as anodes.

### Behavioural outcome variables

#### GRASSP

The primary behavioural outcome of interest was hand function as assessed by the GRASSP[16]. The GRASSP is a battery of clinical outcome tests developed to assess upper extremity function in individuals with tetraplegia and consists of 5 subtests: Manual Muscle Testing (MMT), in which range of motion and strength are assessed in ten muscles on both arms, Semmes and Weinstein Monofilament (SWM), in which touch sensation is measured with microfilaments of increased width across 3 dorsal (D sensation) and 3 palmar (P sensation) locations for each hand; Qualitative Prehension, which assesses the ability to reproduce three grasp patterns (cylindrical grasp, lateral key pinch, and tip to tip pinch); Quantitative Prehension, in which 6 prehension tasks (pouring water from a bottle into a jar, unscrewing lids from two jars, moving pegs into a pegboard, using a key, inserting coins into an aperture, placing nuts onto screws) are completed.

#### Spinal Cord Independence Measure III (SCIM III)

Independence was assessed with the Spinal Cord Independence Measure III (SCIM III) questionnaire[17]. The SCIM III is an independence scale specifically developed to assess independence in people living with spinal cord injury. The SCIM III consists of three subscales: self-care, respiration and sphincter management and (3) mobility.

#### Quality of Life (QLI-SCI)

Quality of life was assessed with the SCI version of the Quality of Life Index (QLI-SCI)[18], a widely used test to assess quality of life in people living with spinal cord injury. It consists of 37 items covering aspects of: Health and Functioning; Social and Economic Subscale; Psychological/Spiritual Subscale; Family Subscale.

#### Grasping and lifting task

During the functional assessment sessions, participants were asked to complete standardised unimanual grasping and lifting movements. Two custom-built manipulanda were used throughout the tasks. Each manipulandum weighted 400 g, contained a 50 N load cell (Make, model: Omega, LCM201-50) with grip force data processed using a 16-bit data acquisition card (National Instruments, USB-6002) and a custom-built program in Labview (v.14)[19]. Three unimanual movements for each hand were performed in a blocked, randomised order. At the beginning of the task, manipulanda were placed at a comfortable starting position for each participant (∼20 cm on average) on the horizonal plane and parallel with their shoulders. Instructions to the participants were to grasp the manipulandum with the required hand, raise them until shoulder height or as close to it as achievable by the participant and move them as far as possible on the horizontal plane (e.g., away from the body by extending the arms). Force data were collected throughout the movements at a sampling frequency of 200 Hz.

### Neurophysiological outcome variables

#### Surface Electromyography (EMG)

Surface EMG activity was recorded bilaterally from the following muscles and positions: abductor pollicis brevis (APB), with the recording electrode at the midpoint between the first metacarpophalangeal joint and carpometacarpal joint[20]; flexor carpi radialis (FCR), with the recording electrode at one-third of the distance from the medial epicondyle to radial styloid[21]; extensor carpi radialis longus (ECRL), with the recording electrode at one-sixth of the distance from the lateral epicondyle to radial styloid[22]. EMG activity was recorded using parallel-bar wireless sensors (3.7[×[2.6 cm) (Trigno, Delsys Inc., Natick, MA, USA) for FCR and ECR and a parallel-bar wireless mini sensor (Trigno, Delsys Inc., Natick, MA, USA) for APB. EMG traces were pre-amplified (gain = 909) with a 20-450 Hz bandwidth and digitized at 2 kHz with the Spike2 (Cambridge electronics Design, Cambridge, UK) data acquisition software[23].

#### TMS Motor-evoked potentials (MEPs)

Magnetic stimulation was applied to the left primary motor area (M1) via a Magstim BiStim² stimulator and a flat alpha coil (Magstim Company, Whitland, Dyfed, UK) being held by a support stand (Magstim Company, Whitland, Dyfed, UK). Stimuli were delivered at a rate of 0.2 Hz while participants were wearing sound-attenuating headphones in order to reduce the effects of sound on the excitability of the corticospinal tract and increase the validity of MEPs as a measure of corticospinal excitability[24]. The coil was oriented at ∼45° to induce a posterior-to-anterior current flow perpendicular to the central sulcus[25]. The optimal coil position to evoke MEPs in APB was found by moving the coil over the scalp while delivering stimulation and marking the position at which MEPs could be elicited at the lowest stimulation intensity to ensure stability of recordings over the session. The position and orientation of the coil was monitored continuously, and if necessary, adjusted to align with the scalp markings[23]. During all the interventions, stimulus delivery was automated through Spike2 (Cambridge Electronic Design, Cambridge, UK). Resting motor threshold (MT) was determined through the relative frequency method[26] by identifying the smallest intensity of stimulation (in % of maximal stimulator output, MSO) necessary to elicit peak-to-peak MEP amplitudes between 50 and 100 μV in at least 5 out of 10 trials in the APB muscle, plus 1. Once the MT was estimated, MEP recruitment curves were obtained by delivering TMS at 10% increments of intensity between 90 and 150% of the MT, with 10 stimuli delivered at each level of intensity for each time, participant, and session[27].

#### TCES spinal-evoked potentials

Single-pulse TCES was delivered through a Digitimer DS8R Constant Current Stimulator (Digitimer, Welwyn Garden City, Hertfordshire, UK) controlled by the Spike2 (Cambridge Electronic Design, Cambridge, UK) software. Stimulation was delivered through a pair of self-adhesive electrodes (Axelgaard, ValuTrode Cloth): a 5×9 cm electrode was placed over the left iliac crest as anode; a 3.2 cm round electrode was placed at the midline between C5 and C6 spinous processes as cathode[11]. In order to elicit spinal responses, TCES pulses were delivered using 1ms biphasic square-wave pulses delivered every 5 seconds[28]. Spinal-evoked potentials were recorded bilaterally from the APB, FCR and ECR muscles. Spinal-evoked threshold was determined for each participant and session as the lowest intensity of stimulation at which spinal responses of amplitudes >50 μV could be observed in each of the three muscles[29]. Once the threshold value was estimated, spinal-evoked potentials recruitment curves were obtained by delivering TCES at 10% increments of intensity between 90 and 150% of threshold, with 10 stimuli delivered at each level of intensity for each time, participant, and session. Ten recordings were obtained at each intensity of stimulation[11]. Upon every increase of intensity, participants were asked to rate their perceived pain level from 0 to 10 using a visual analogue scale for pain, with 0 defined as “no discomfort at all” and 10 as “unbearable pain”. Stimulation was halted immediately if discomfort reached level 8 out of 10[12]. No participants reported discomfort levels higher than 6 out of 10 in any of the sessions.

### Data analyses

For all variables, data collected during the two baseline sessions were averaged to compute a single baseline value.

#### Functional outcome variables

For the analysis of GRASSP data, overall scores for each hand assessed in each session were included in the analysis. A linear mixed-effects model fit by maximum likelihood was run (SPSS software; Version 26.0) with an a priori significance level of 0.05. Participant was included as a random factor, with Time (Baseline, Post-1, Post-2, Follow-up), Order (ULTP first, ULTP+TCES first) and Hand (Right, Left) as fixed factors. In order to determine whether the observed effects are clinically relevant, we measured how many participants met or exceeded the minimally detectable difference (MDD) and Minimal clinically important difference (MCID) criteria for the strength, sensation and quantitative prehension subdomains[30, 31].

For the analysis of SCIM III and QLI-SCI data, overall scores calculated in each session were included in the analysis. A linear mixed-effects model fit by maximum likelihood was run (SPSS software; Version 26.0) with an a priori significance level of 0.05. Participant was included as a random factor, with Time (Baseline, Post-1, Post-2, Follow-up) and Order (ULTP first, ULTP+TCES first) and Hand (Right, Left) as fixed factors.

For force data during the unimanual grasp and lifting tasks, we first identified the onset of each trial as the first timepoint for which the force produced in the relevant hand exceeded 10% of the peak force value collected throughout all trials, and the offset of each trial as the first timepoint for which the force produced in the relevant hand returned to values below 10% of the peak force value collected throughout all trials. This preliminary data extraction strategy was carried out to exclude from our analysis all datapoints during which the participants were not grasping the manipulandum. We then computed a single mean grip force value for the three movements completed with the same hand in every session. We ran linear mixed-effects model fit by maximum likelihood (SPSS software; Version 26.0) with an a priori significance level of 0.05 on the average force values. Since force values were strongly influenced by the hand function (Supplementary Fig. 2), we modelled the effect of Dominance by computing a binary variable (1 = Right, 2 = Left) and identifying participants’ most functional hand based on their GRASSP scores. Participant was included as a random factor, with Time (Baseline, Post-1, Post-2, Follow-up), Order (ULTP first, ULTP+TCES first), Dominance (Right, Left) as fixed factors.

#### Neural outcome variables

Changes in corticospinal excitability were assessed by comparing MEPs amplitudes collected upon TMS at each time point. We calculated the peak-to-peak amplitude for each MEP and averaged the 10 MEPs recorded at each intensity of stimulation from 90%MT to 150% MT. In two participants (P618, P650), we could not induce any reliable MEPs in the APB muscle: thresholding and recruitment curves were therefore based on the FCR muscle in these participants. A linear mixed-effects model fit by maximum likelihood was run (SPSS software; Version 26.0) with an a priori significance level of 0.05. Participant was included as a random factor, with Time (Baseline, Post-1, Post-2, Follow-up), Order (ULTP first, ULTP+TCES first) and Intensity (90%MT, 100%MT, 110%MT, 120%MT, 130%MT, 140%MT, 150%MT) as fixed factors.

Changes in spinal excitability were assessed by comparing spinal-evoked potential amplitudes collected upon single-pulse TCES at each time point. For the TCES data, we calculated the peak-to-peak amplitude for each spinal-evoked potential response recorded from the three muscles of interest (APB, FCR, ECR) from each arm and averaged the 10 spinal responses recorded at each intensity of stimulation from 90% to 150% of threshold. A linear mixed-effects model fit by maximum likelihood was run (SPSS software; Version 26.0) with an a priori significance level of 0.05. Participant was included as a random factor, with Time (Baseline, Post-1, Post-2, Follow-up), Order (ULTP first, ULTP+TCES first), Intensity (90% threshold, 100% threshold, 110% threshold, 120% threshold, 130% threshold, 140% threshold, 150% threshold) and Hand (Left, Right) as fixed factors.

Effect sizes were interpreted as follows: η*_p_*^2^<0.01 indicates a small effect; 0.06< η*_p_*^2^<0.14 indicates a medium effect; η_p_ ^2^>0.14 indicates a large effect[32]. Importantly, because of the inclusion of random effects in the analysis, only partial eta squared, which estimate the proportion of variance explained by a given parameter relative to the variance in a model without that parameter, could be estimated[33].

## Results

### GRASSP

The linear mixed-effects analysis revealed a significant Time*Order interaction on the GRASSP scores [*F* (3, 17)=5.313, *p*=0.009, η*^2^*=0.48]. For Group_1, pairwise comparisons showed a significant increase in GRASSP scores from Baseline to Post-2 (*p<*0.001), from Post-1 to Post-2 (*p<*0.001) and from Baseline to Follow-up (*p=*0.017) (Figure 2, *A*). For Group_2, pairwise comparisons showed a significant increase in GRASSP scores from Baseline to Post-1 (*p<*0.001), from Baseline to Post-2 (*p<*0.001) and from Baseline to Follow-up (*p<*0.001) (Figure 2, *B*). All the other interactions and main effects are reported in Table 2. Scores for each of the five (strength, dorsal sensation, palmar sensation, qualitative prehension, quantitative prehension) are reported in Supplementary Fig. 3. This indicates that ULTP+TCES significantly increase hand function and that the increases persisted for three months after the interventions.

**Table 2.** Fixed-effects table for the linear mixed model run on the GRASSP scores. η_p_ ^2^ = partial eta squared.

| Parameter | Numerator df | Denominator df | F | Sig. | $\eta_p^2$ |
| --- | --- | --- | --- | --- | --- |
| Time | 3 | 17 | 13.243 | <0.001 | 0.70 |
| Hand | 1 | 7 | 0.433 | 0.531 | 0.06 |
| Order | 1 | 7 | 0.005 | 0.947 | <0.001 |
| Time*Hand | 3 | 17 | 0.804 | 0.509 | 0.12 |
| Time*Order | 3 | 17 | 5.313 | 0.009 | 0.48 |
| Hand*Order | 1 | 7 | 0.902 | 0.373 | 0.11 |
| Time*Hand*Order | 3 | 17 | 1.130 | 0.365 | 0.17 |

**Figure 2.**
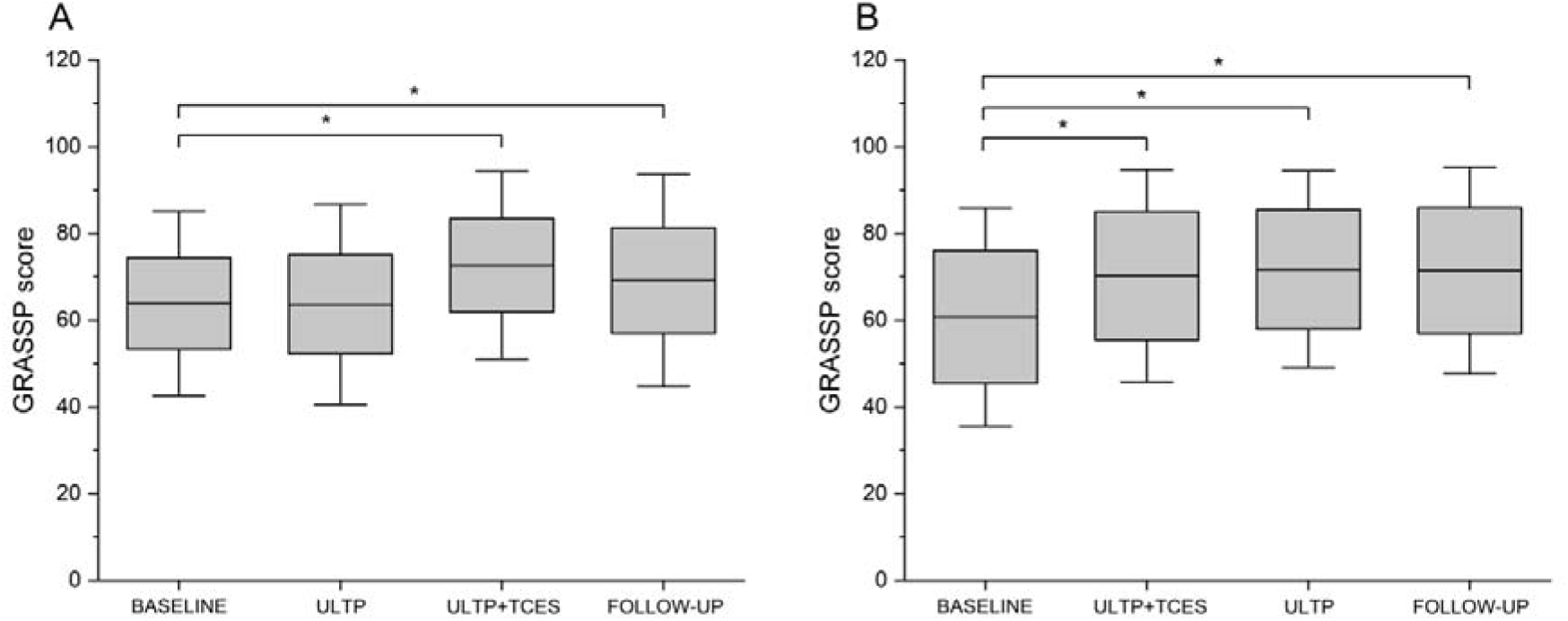
Total GRASSP scores across the functional assessment sessions for participants assigned to Group_1 *(A)* and Group_2 *(B).* Asterisks indicate significant differences; boxes represent the associated standard error (SE) and whiskers represent the associated 95% confidence interval.

Aside from the significant effect observed at the group level, we report that GRASSP scores exceeded the minimally detectable difference (MDD) criteria[30]: for strength, in 4 participants out of 5 after ULTP+TCES and in 3 participants out of 5 at follow-up; for sensation, in 3 participants out of 5 after ULTP+TCES and in 1 participants out of 5 at follow-up; for quantitative prehension, in 1 participant out of 5 after ULTP+TCES and in no participant at follow-up (Kalsi-Ryan, 2016). GRASSP scores exceeded the Minimum Clinically Important Difference Values (MCID)[31]: for strength, in 1 participant out of 5 after ULTP+TCES and at follow-up; for quantitative prehension, in 3 participants out of 5 after ULTP+TCES and in 3 participants out of 5 at follow-up (Supplementary Table 1).

### SCIM III

The linear mixed-effects analysis revealed no significant interaction between Time*Order interaction on the SCIM III scores [*F* (3, 9)=1.504, *p*=0.279, η*^2^*=0.33] nor significant main effects of Time [*F* (3, 9)=1.438, *p*=0.295, η*^2^*=0.32] and Order [*F* (1, 9)=0.699, *p*=0.464, η =0.19] (Supplementary Table 2).

Analysis by subscales further confirmed the lack of significant effects for self-care (Time*Order interaction, *p*=0.191), respiration and sphincter management (Time*Order interaction, *p*=0.476) and mobility (Time*Order interaction, *p*=0.659). This suggests that none of the interventions affected independence.

### QLI-SCI

The linear mixed-effects analysis revealed a significant Time*Order interaction on the QLI-SCI scores [*F* (3, 9)=6.787, *p*=0.011, η*^2^*=0.69]. For Group_1, pairwise comparisons showed a significant increase in QLI-SCI scores from Baseline to Post-1 (*p=*0.002), from Baseline to Post-2 (*p<*0.001) and from Baseline to Follow-up (*p=*0.013). For Group_2, pairwise comparisons showed no significance difference between QLI-SCI scores collected at any timepoint (Baseline-Post1, *p*=0.289; Baseline-Post2, *p*[=0.609; Baseline-Follow-up, *p*=0.893). The main effect of Time was significant [*F* (3, 9)=5.817, *p*=0.017, η*^2^*=0.66] and the main effect of Order was non-significant [*F* (1, 9)=0.092, *p*=0.781, η*^2^*=0.01] (Supplementary Table 2). Analysis by subscales showed no significant effects for Health and Functioning (Time*Order interaction, *p*=0.108), Social and Economic (Time*Order interaction, *p*=0.597), Psychological/Spiritual (Time*Order interaction, *p*=0.057) and Family (Time*Order interaction, *p*=0.652) subscales. This indicates that both interventions increased quality of life for participants in Group_1 and that the increases persisted for three months after the interventions.

### Grasping and lifting task

The linear mixed-effects analysis run on the average grip force values revealed a significant three-way interaction of Time*Order*Dominance [*F* (3, 6)=7.641, *p*=0.018, η*^2^*=0. 79]. For Group_1, pairwise comparisons showed no significant changes in force in the dominant hand from Baseline to Post-1 (*p*=0.142), from Baseline to Post-2 (*p*=0.951) nor from Baseline to Follow-up (*p*=0.053), but changes in force in the non-dominant hand from Baseline to Post-2 (*p*=0.027) (Supplementary Fig. 4, *A*). For Group_2, pairwise comparisons showed a significant increase in force in the dominant hand from Baseline to Post-1 (*p*<0.001), and from Baseline to Follow-up (*p*=0.002) (Supplementary Fig. 4, *B*), but no significant increases in force in the non-dominant hand from Baseline to Post-1 (*p*=0.757), Baseline to Post-2 (*p* =0.436) or Baseline to Follow-up (*p*=0.234). All the other interactions and main effects are reported in Supplementary Table 3. This suggests that ULTP+TCES increases grip force and, for group 2, this increase persisted for three months after the interventions.

### TMS MEPs

Mean and *SD* of the MT values across all participants were 48±2 % MSO. The linear mixed-effects analysis run on the MEPs at MT intensity revealed a significant interaction between Time*Order [*F* (3, 63)=6.260, *p*<0.001, η*^2^*=0.23]. For Group 1, pairwise comparisons showed no significant increase in MEP amplitudes from Baseline to Post-1 (*p*=0.06), from Baseline to Post-2 (*p*=0.969) nor from Baseline to Follow-up (*p*=0.735) (Figure 3, *A*). For Group_2, pairwise comparisons showed a significant increase in MEP amplitudes from Baseline to Post-2 (*p*=0.013) and from Post-1 to Post-2 (*p*<0.001), but no significant changes from Baseline to Post-1 (*p*=0.153) nor from Baseline to Follow-up (*p*=0.989) (Figure 3, *B*). All the other interactions and main effects are reported in Table 3. This suggests that, for participants in Group 2, ULTP significantly increased corticospinal excitability but the effects did not persist at three months after the interventions.

**Figure 3.**
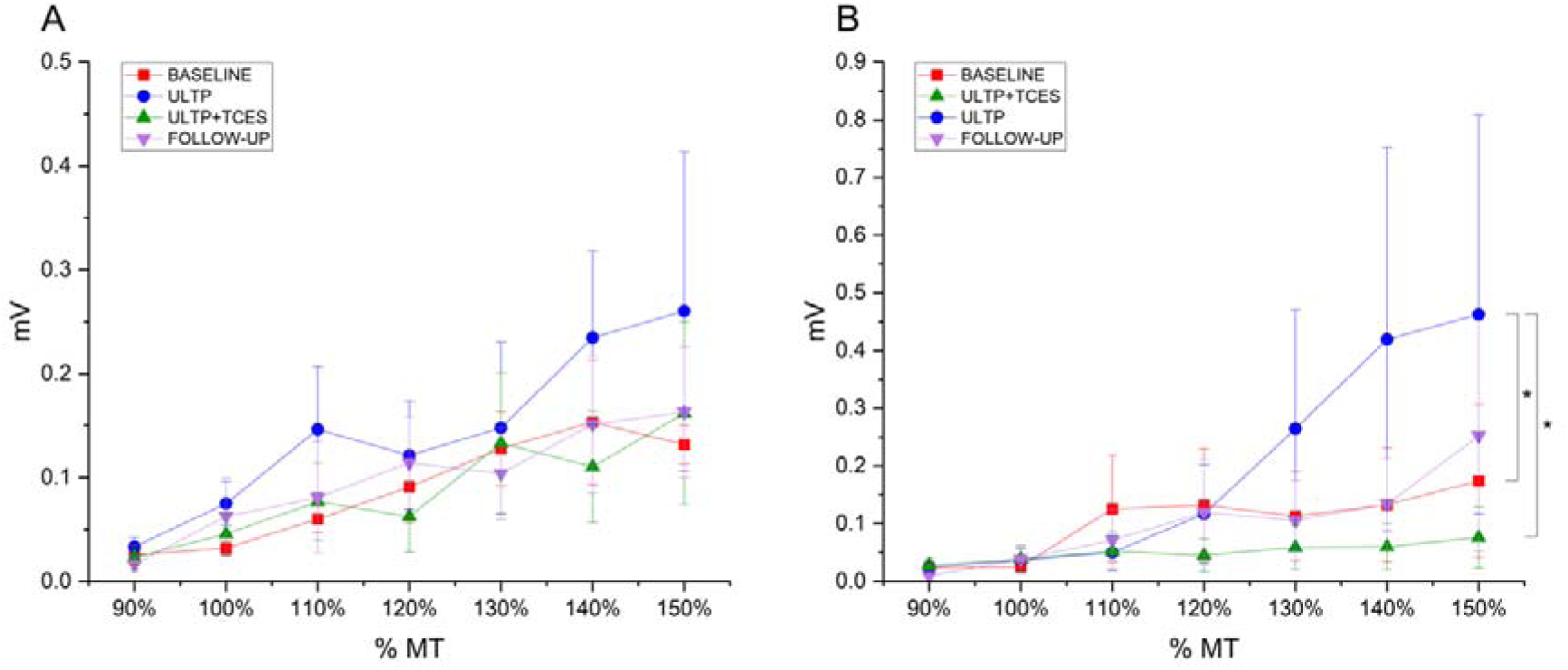
MEP recruitment curves across the functional assessment sessions for participants assigned to Group_1 *(A)* and Group_2 *(B).* Asterisks indicate significant differences and whiskers represent the associated standard error (SE).

**Table 3.** Fixed-effects table for the linear mixed model run on the MEP amplitude values. η*_p_*^2^ = partial eta squared.

| Parameter | Numerator df | Denominator df | F | Sig. | $\eta_p^2$ |
| --- | --- | --- | --- | --- | --- |
| Time | 3 | 63 | 1.714 | 0.173 | 0.08 |
| Intensity | 6 | 21 | 1.610 | 0.194 | 0.32 |
| Order | 1 | 21 | 0.048 | 0.828 | <0.001 |
| Time* Order | 3 | 63 | 6.260 | <0.001 | 0.23 |
| Time*Intensity | 18 | 63 | 0.717 | 0.782 | 0.17 |
| Intensity*Order | 6 | 21 | 0.068 | 0.998 | 0.19 |
| Time*Order*Intensity | 18 | 63 | 1.064 | 0.407 | 0.21 |

### TCES spinal-evoked potentials

Mean and *SD* of the threshold intensity for TCES across all participants were 34±6 mA. The linear mixed-effects analysis run on the spinal evoked responses revealed a significant three-way interaction of Time*Order*Hand [*F* (3, 193)=7.073, *p*<0.001, η*^2^*=0.10]. For Group_1, pairwise comparisons showed a significant increase in amplitude of the left arm from Baseline to Post-2 (*p*<0.001) (Figure 4, *A*) but no significant changes from Baseline to Post-1 (*p*=0.443), or from Baseline to Follow-up (*p*=0.333) (Figure 4, *C*). For Group_2, pairwise comparisons showed no significant change in amplitude of the left arm from Baseline to Post-1 (*p*=0.476), Baseline to Post-2 (*p*=0.381) or from Baseline to Follow-up (*p*=0.109) (Figure 4, *B*), but significant increases in amplitude of the right arm from Baseline to Post-1 (*p*<0.001) and from Baseline to Follow-up (*p*=0.04) (Figure 4, *D*). All the other interactions and main effects are reported in Table 4. This suggests that ULTP+TCES increased spinal excitability and, for participants in Group 2, this increases persisted for three months after the end of the interventions.

**Table 4.** Fixed-effects table for the linear mixed model run on the spinal-evoked potentials amplitude values. η*_p_*^2^ = partial eta squared.

| Parameter | Numerator df | Denominator df | *F* | Sig. | $\eta_p^2$ |
| --- | --- | --- | --- | --- | --- |
| Time | 3 | 175 | 2.915 | 0.036 | 0.05 |
| Intensity | 6 | 175 | 11.042 | <0.001 | 0.27 |
| Order | 1 | 175 | 0.120 | 0.730 | <0.001 |
| Hand | 1 | 175 | 0.250 | 0.618 | <0.001 |
| Time*Intensity | 18 | 175 | 0.418 | 0.983 | 0.04 |
| Order*Intensity | 6 | 175 | 0.075 | 0.998 | <0.001 |
| Hand*Intensity | 6 | 175 | 0.189 | 0.980 | <0.001 |
| Time*Order | 3 | 175 | 12.907 | <0.001 | 0.18 |
| Time*Hand | 3 | 175 | 1.595 | 0.192 | 0.03 |
| Order*Hand | 1 | 175 | 3.370 | 0.068 | 0.02 |
| Time*Order*Hand | 3 | 175 | 7.073 | <0.001 | 0.11 |
| Intensity*Order*Hand | 6 | 175 | 0.108 | 0.995 | <0.001 |
| Intensity*Time*Order | 18 | 175 | 1.402 | 0.135 | 0.13 |
| Intensity*Time*Hand | 18 | 175 | 0.199 | 1.00 | 0.02 |
| Intensity*Time*Hand*Order | 18 | 175 | 0.359 | 0.993 | 0.04 |

**Figure 4.**
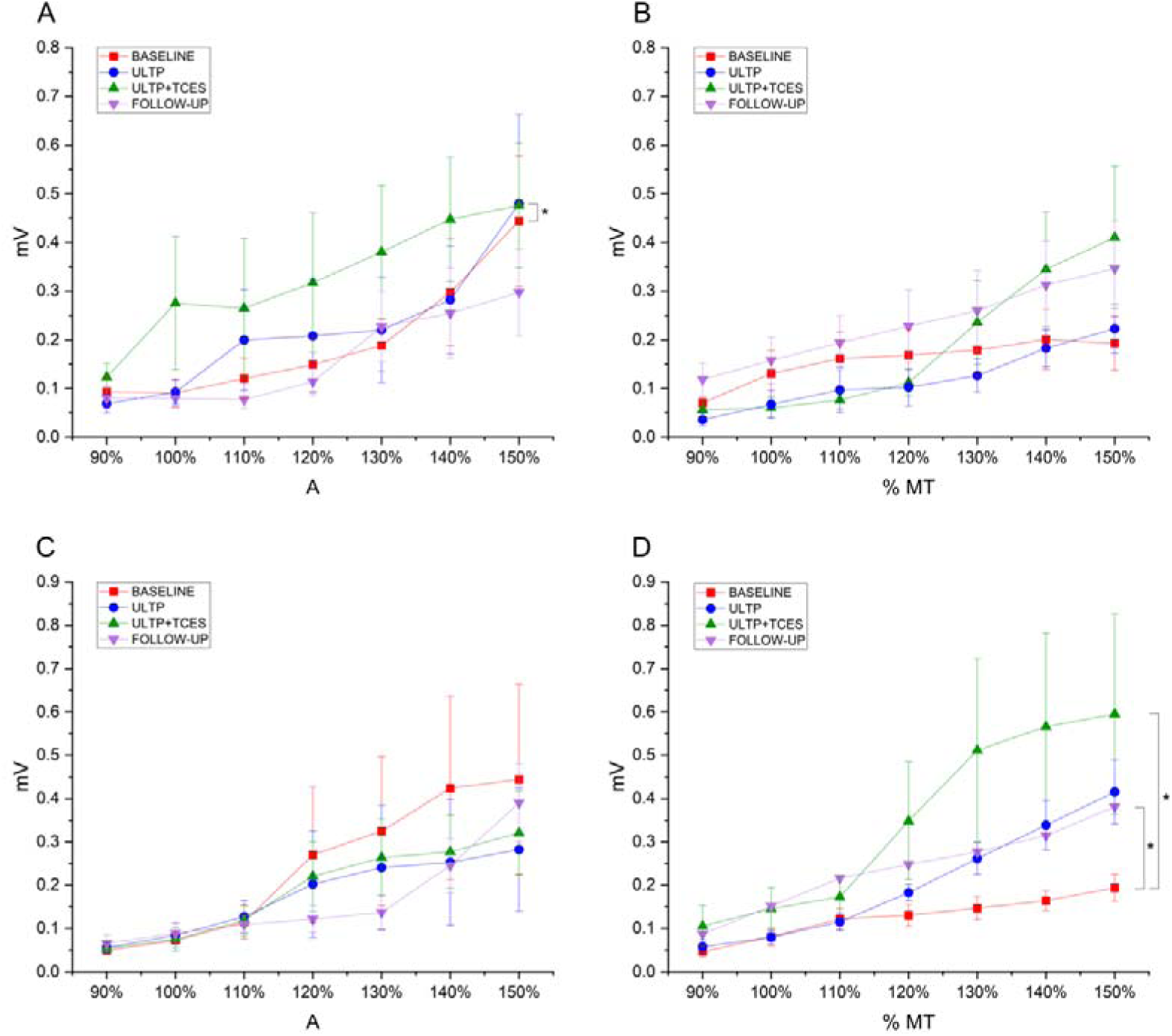
Spinal-evoked potentials recruitment curves collected from the left arm across the functional assessment sessions for participants assigned to Group_1 *(A)* and Group_2 *(B).* Spinal-evoked potentials recruitment curves collected from the right arm across the functional assessment sessions for participants assigned to Group_1 *(C)* and Group_2 *(D).* Asterisks indicate significant differences and whiskers represent the associated standard error (SE).

## Discussion

The aims of the current study were to: (1) compare the effects of one month of ULTP followed by one month of ULTP+TCES with the effects of one month of ULTP+TCES followed by one month of ULTP on hand function, independence and quality of life; (2) assess changes in corticospinal excitability occurring throughout the intervention and at three/months after the end of the intervention; (3) assess changes in spinal excitability occurring throughout the intervention and at three/months after the end of the intervention; (4) determine if the observed effects on hand function are paralleled by changes in force production while participants complete grasping and lifting tasks.

### Functional outcomes

Participants were assigned to either Group_1, for which they completed a month of ULTP followed by a month of ULTP+TCES, or Group_2, for which they completed a month of ULTP+TCES followed by a month of ULTP. For participants in Group_1, significant improvements in hand function were only observed after the combinatorial intervention. In contrast, for participants in Group_2, a month of ULTP+TCES was sufficient to induce significant hand function improvements (see Figure 2). Similar results were reported by Inanici et al.[7] who observed higher cumulative improvements in GRASSP scores after functional task training paired with spinal stimulation compared with the ones observed after functional task training. Nevertheless, the authors also reported significant changes in GRASSP scores from baseline to after functional task training[7], changes which were not evident in the current study. This discrepancy could be explained by the different methods employed in the two studies: first, the intensive task practice employed by Inanici et al.[7] consisted of two hours of activity per session while the ULTP protocol we employed only lasted for about 30-40 minutes per session; participants in the Inanici et al.[7] study received a total of 24 task practice sessions over two months, twice as much as the participants involved in our study. Taken together, these findings suggest that the frequency and length of the ULTP (delivered in isolation) used in our study was not sufficient to increase manual performance and that the combined effect of the two interventions is hypothesised to be the cause of the improvement in manual performance. Our data make an important contribution to the literature suggesting that task practice and stimulation have additive effects on function as observed in animal models[34].

Interestingly, no improvements were observed in GRASSP scores after a further month of ULTP following one month of ULTP+TCES. This is consistent with the literature showing that people living with SCI might reach a plateau in functional recovery after practice[4, 35] and suggests that combinatorial strategies are necessary to improve motor function[36]. Whether the plateau we observed could be overcome with further stimulation remains a matter of dispute. Finally, the persistent (three months after the last intervention session) improvements in hand function observed in both participants’ groups constitute a promising approach for long-term rehabilitation, without the need for stimulation to be delivered continuously while other rehabilitation strategies are explored to maintain the motor system in a more functional state.

In order to assess the clinical relevance of the observed functional improvements, we measured whether GRASSP scores measured at each time point met or exceeded the criteria for minimally detectable difference (MDD) and Minimal clinically important difference (MCID) for the strength, sensation and quantitative prehension subdomains. Clinically important changes were observed in 1 participant out of 5 after ULTP+TCES and at follow-up for strength, and 3 participant out of 5 after ULTP+TCES and at follow-up for prehension (Table 3). Importantly, the values we based the analysis on were computed values by assessing spontaneous changes occurring longitudinally over the first six months after injury [31], time range at which functional recovery is greatest [37]. This suggests that spinal stimulation can contribute to changes in hand function similar in magnitude to the ones spontaneously observed in the acute phase after SCI even after many years from the injury (range 2-30 years after injury in the current study). Clinically important changes in prehension were observed even in one participant who has been living with SCI for the last 30 years (P528).

In addition, we explored whether changes in upper limb function were paralleled by perceived changes in independence and quality of life, as assessed through self-reported questionnaires (SCIM III for independence, QLI-SCI for quality of life). No significant changes from baseline in independence were observed after either of the interventions or at follow-up. A previous study[7] in which a similar cross-over design was employed to assess the efficacy of spinal stimulation and training reported improvements between 1 and 4 points in the SCIM self-care domain, results which are in line with the current study (Supplementary Table 1). However, Inanici et al.[7] did not report any statistical analysis on the SCIM III scores. Changes observed after rehabilitation alone and rehabilitation paired spinal stimulation by Moritz et al.[8] failed to reach statistical significance. Interestingly, our results suggest that quality of life significantly increased for participants in Group_1 (Supplementary Table 2). already after one month of ULTP, and improvements were maintained throughout the study and follow-up. Nevertheless, no significant changes were observed in participants assigned to Group_2 at any time points. The use of instruments to estimate quality of life after SCI have been criticised as they underestimate the impact of factors unrelated to health (e.g. the psychological/spiritual and family subscales of the QLI-SCI)[38]. As we observed improvements in scores for items whose change are unlikely due to the intervention employed (e.g. a 5-points increase in the score of the Psychological/Spiritual subscale and 5-points increase in the score of the Family subscale), we argue that these non-conclusive results observed could be explained by factors not directly relevant to study participation and call for further studies specifically assessing changes in quality of life attributable to spinal stimulation in people living with SCI.

We ought to investigate whether the improvements observed with the GRASSP also translated to a different motor task in which participants were asked to grasp and lift manipulanda containing load cells that could objectively measure the grip force produced. Results from this task closely matched the ones observed for GRASSP and spinal-evoked potentials data, in that a month of ULTP paired with spinal stimulation increased hand function, the average force produced during grasping and lifting trials and increased spinal excitability. In addition, for participants in Group_2, persistent increases in GRASSP score and grasping/lifting forces were still evident at 3-months follow-up and paralleled by persistent higher spinal excitability. However, and to our surprise, these effects were also proved to depend on hand dominance. For participants in Group_1 increases in average force were only observed after ULTP+TCES in the non-dominant hand, while for participants in Group_2 increases in average force were observed after ULTP+TCES and at follow-up in the non-dominant hand. Gad et al.[13] demonstrated that cervical stimulation increased maximum grip strength in all participants irrespective of hand dominance. We speculate that different levels of grip forces at baseline might explain the lack of improvements observed in the non-dominant in Group_1: participants 192 and 618 exhibited very limited strength (all muscles distal to ECR graded 0-1 for Manual Muscle Testing) and prehension (graded 1 for qualitative prehensive). This hypothesis is in line with the evidence that TCES necessitates of a minimal level of handgrip force to promote functional improvements[39]. This issue also underlines the need for stratification of neuromodulatory interventions based on the capabilities and needs of each individual. Our findings indicate that behavioural improvement can be achieved through combinatorial rehabilitation practices[40].

### Neural outcomes

We assessed changes in corticospinal excitability by delivering TMS at intensities ranging from 90% to 150% of motor threshold throughout the study. First, we demonstrated that completing a month of ULTP after one month of ULTP+TCES increased the amplitude of TMS-evoked responses. Together with the trend towards significance (p = 0.06, Cohen’s d = 0.4 indicating a small to medium effect size) observed for Group_1 after ULTP, our findings add to the body of literature showing that task practice induced neural plasticity along the corticospinal tract[41, 42]. Nevertheless, we also showed that the increase in the excitability was not sustained at follow up three-months after the end of the intervention, despite the persistent increase in function at this late stage. These findings have two important implications: firstly, we suggest that whichever neural populations were upregulated by the ULTP, their excitability returned to baseline values after a period without further task practice. This interpretation is in line with the evidence accumulated from motor skill learning studies showing how the neural networks involved in learning progress from MI-SMA during the initial phases towards a more distributed network involving also cerebellar and striatal structures[43]. Second, that plasticity along the corticospinal tract is not determinant in inducing long-lasting functional changes after SCI, since no significant change in function was observed after ULTP alone and no increases in MEP amplitudes were observed after ULTP+TCES. Regarding this latter point, recent evidence suggests that the effects of spinal stimulation on cortical structures as measured through TMS might depend on the stimulation frequency employed: for example, Murray & Knikou[44] reported that 14 sessions of spinal stimulation delivered at low frequency (0.2 Hz) significantly increased MEP amplitudes. In contrast, Benavides et al. [11]further investigated how frequency affects corticospinal output by showing that MEP amplitudes increased when spinal stimulation was applied with 30Hz pulses without a 5 kHz carrier frequency but did not change when the 5 kHz carrier frequency was employed after a single session. As we similarly employed a carrier frequency of 10kHz, it is plausible that this high-frequency component had an inhibitory effect on cortical excitability which counteracted the excitatory effects of ULTP. Further investigation is needed to characterise the impact of stimulation frequencies on the induction of cortical plasticity and on upper limb function.

Spinal excitability was assessed by stimulating the spinal cord at the C5-C6 level with threshold and suprathreshold intensities. For participants in Group_1, adding spinal stimulation after a month of ULTP significantly increases spinal excitability measured from the left arm (most affected arm in two of the three participants). Similar effects, although in the right arm, were observed for Participants in Group_2 after one month of ULTP+TCES. These findings are largely in agreement with studies investigating the acute[11] and longer-term[7, 13, 14] effects of spinal stimulation on spinal circuitry, all of which reported increases in spinal excitability after cervical non-invasive stimulation. Interestingly, our study is the first to our knowledge to report different effects of stimulation on spinal excitability measured from the left and right arm muscles. A possible explanation lies in the difference in stimulation loci employed: while all previous studies positioned the electrodes at the midline, we selectively targeted the more affected arm through a mapping procedure identifying which position elicited the highest responses in the most affected arm[45]. Finally, the divergence between the GRASSP results, indicating bilateral increases in function, and the neural results, indicating no changes in spinal excitability for the left arm in Group_2 and the right arm in Group_1, raises the question on whether other neural mechanisms which cannot be assessed via single-pulse TCES might underlie functional improvements. Importantly, while we personalised TCES according to each participants’ lesion level, spinal-evoked potentials were only recorded upon stimulation at the midline between C5 and C6. Future works further investigating plasticity along multiple spinal segments could potentially extend our findings. Regarding the possible neural mechanism, modulating spinal cord excitability can shift motor network excitability closer to firing threshold, making the ULTP more likely to engage previously inactive neurons[4]. The higher excitability leads to increases in the force produced which constitute the neural basis of the functional improvements observed at longer timescales[46].

### Limitations

There are several limitations to consider in the context of this study. First, we acknowledge the difficulty of drawing conclusions about a clinical population as diverse as spinal cord injury based on a limited number of study participants. Our final sample size was negatively affected by the huge physical burden of committing to travel to our lab for a total of 34 sessions over the course of 25 weeks: of the 27 participants who showed interest in participating and met all the inclusion criteria, only 6 could commit to the full duration of the study and one participant further decided to withdraw from it after the first experimental session. This issue underlines the barriers to trial participation and the need for collaborative multicentre trials to ensure more people have access to potentially beneficial therapies.

Notwithstanding, the five participants who were finally included successfully completed all experimental sessions, resulting in a 100% protocol adherence. Our final sample size is in line with similar works investigating the effects of non-invasive stimulation after SCI[7, 13]. The crossover design we employed additionally renders statistical interpretation problematic, since the carryover effect is inherent to sequential intervention designs[47]. We partially overcame this limitation by stratifying analysis by sequence groups and modelling the effect of Order[47]. Moritz et al.[8], who observed clinically meaningful improvements in arm function when spinal stimulation followed two months of rehabilitation alone, raised the possibility that the initial rehabilitation improved the potential for further effects after spinal stimulation. By employing a crossover design, we observed that one month of spinal stimulation can significantly affect hand function even without any prior rehabilitation. Carryover effects notwithstanding, the analysis show the superiority of pairing ULTP with TCES as opposed to ULTP alone in improving hand function, since GRASSP scores increased from ULTP to ULTP+TCES but not from ULTP+TCES to ULTP.

## Data Availability

The data that support the findings of this study are available from the corresponding author, upon reasonable request.

## Acknowledgements

The author would like to thank Prof. Victor R. Edgerton for constructive criticism of the manuscript.

## Funding

This work was supported by the International Spinal Research Trust.

## Competing interests

Dr Parag Gad has Shareholder interest in SpineX Inc.

## Notes

### Clinical Trial

NCT05801536

### Clinical Protocols

https://clinicaltrials.gov/study/NCT05801536

### Author Declarations

Experimental procedures approved by the HRA and Health and Care Research Wales (HCRW) (REC reference: 22/NW/0171) and conformed to the Declaration of Helsinki. The study was registered with ClinicalTrials.gov (NCT05801536).

### Summary of Updates

Figure 1 missing from original version

