## Supplemental materials for "Bimanual upper limb task practice and Transcutaneous electrical stimulation enhance spinal plasticity and hand function after chronic cervical spinal cord injury"

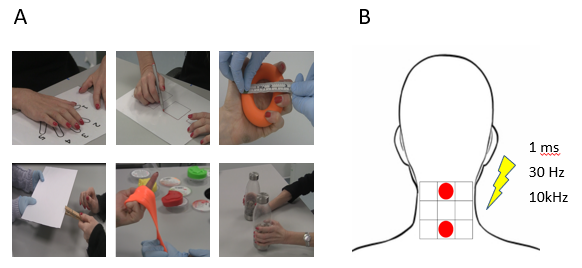

**Supplementary Figure 1.** ULTP comprising of six upper-limb activities of incremental difficulty *(A)*. Five minutes were spent for each activity in each session. Loci and parameters of TCES *(B).*

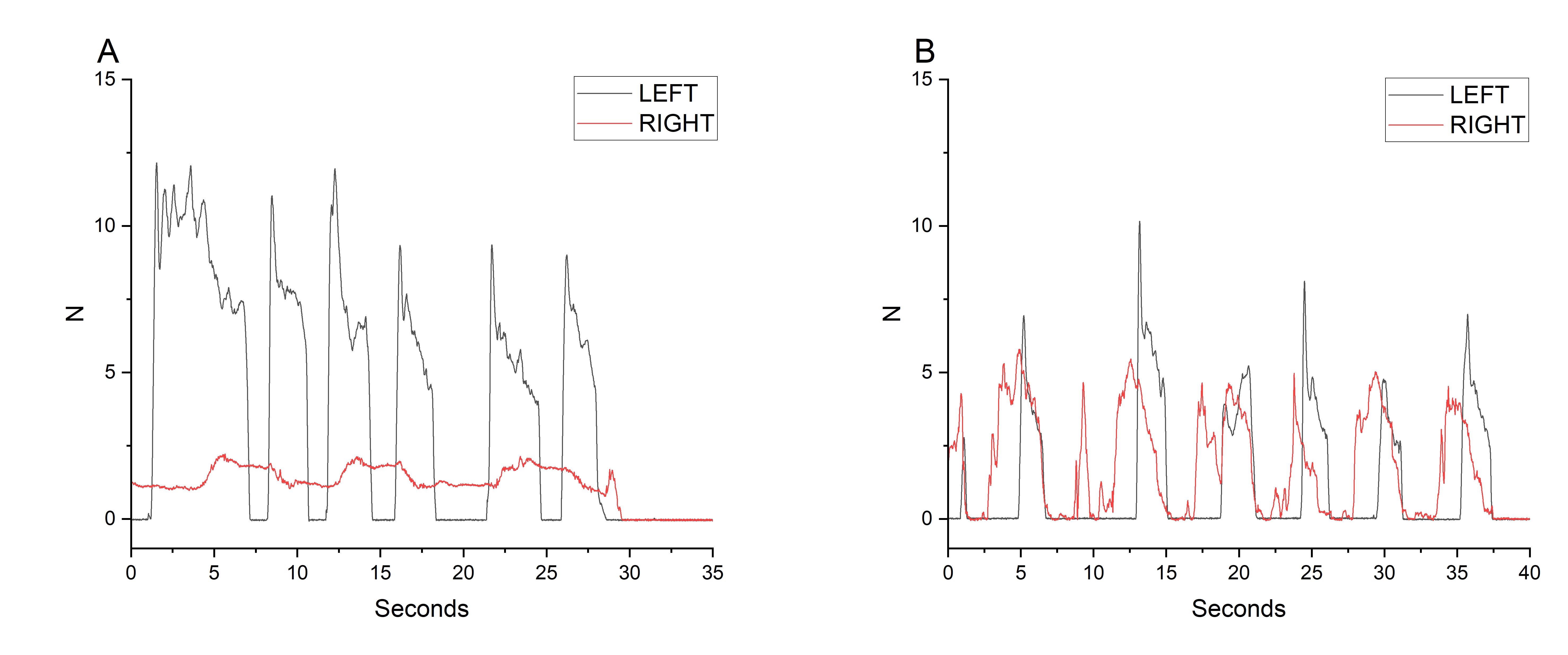

**Supplementary Figure 2.** Grip force (GF)  traces during the grasp-and-lift movements for a representative participant before *(A)* and after one month of ULTP+TCES *(B).*

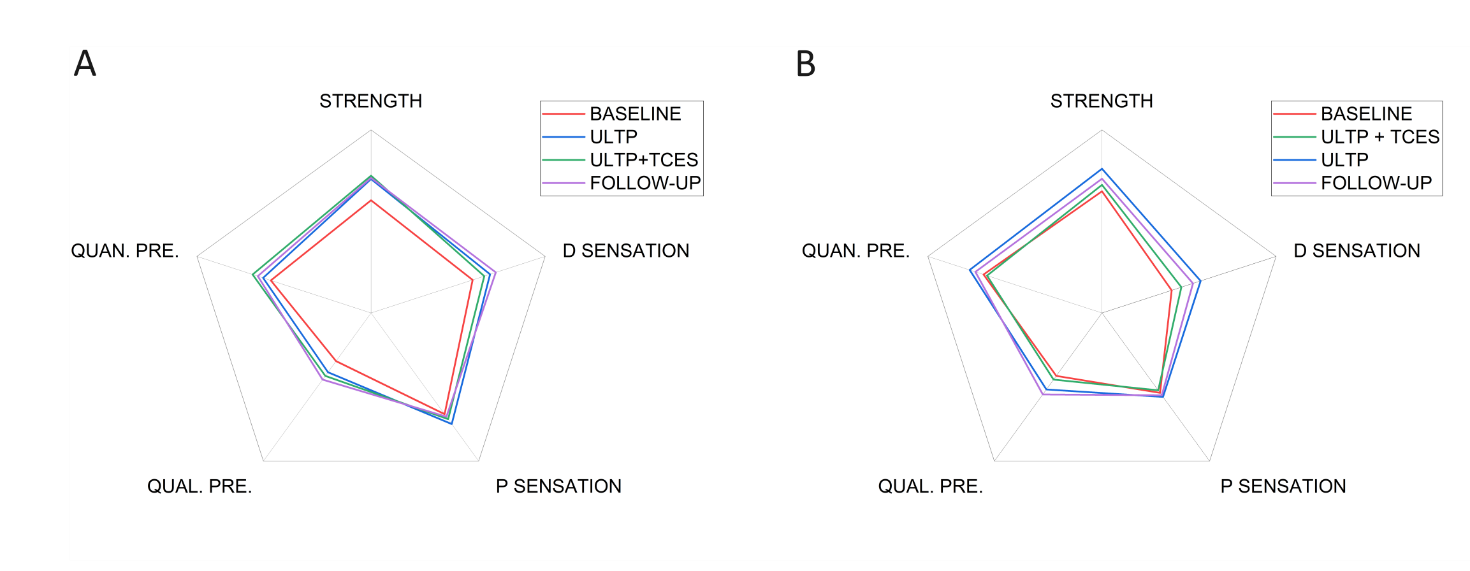

**Supplementary Figure 3.** GRASSP scores throughout the study for Group-1 (*A*, *n* = 3) and Group-2 (B, *n* = 2). Qual. Pre. = qualitative prehension; Quan. Pre. = Quantitative prehension. P sensation = palmar sensation. D sensation = dorsal sensation.

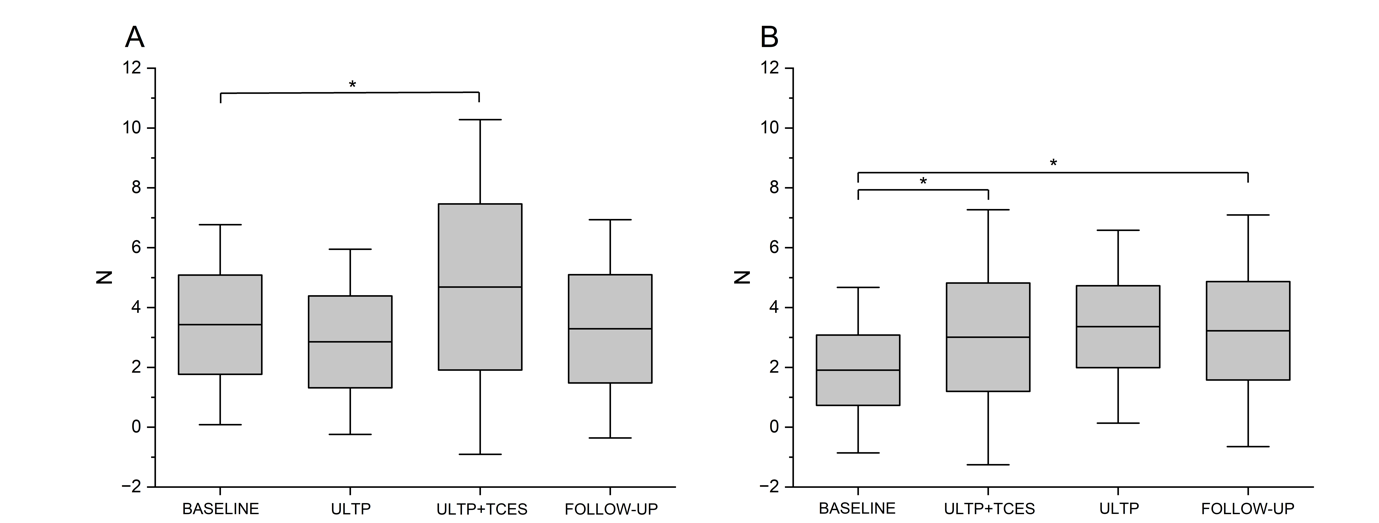

**Supplementary Figure 4.** Average force data collapsed between hands across the functional assessment sessions for participants assigned to Group_1 *(A)* and Group_2 *(B).* Asterisks indicate significant differences, boxes represent the associated standard error (SE) and whiskers represent the associated 95% confidence interval.

**Supplementary Table 1.** Individual GRASSP scores over time by subdomain. MDD = minimally detectable difference (MDD). MCID = Minimal clinically important difference. *Exceeded MDD change. **Exceeded MCID change.

| **Domain** | **Participant** | **Baseline** | **ULTP** | **ULTP+TCES** | **Follow-up** |
| --- | --- | --- | --- | --- | --- |
| *Strength* | 242 | 52.5 | 61* | 67* ** | 67* ** |
| MDD =7 | 192 | 58.5 | 70* | 70* | 71* |
| MCID = 13 | 650 | 39.5 | 41 | 48* | 42 |
|  | 618 | 40 | 47* | 50* | 47* |
|  | 528 | 67.5 | 66 | 74 | 67 |
| *Sensation* | 242 | 29.5 | 27 | 28 | 33 |
| MDD =8 | 192 | 34.5 | 40 | 44* | 39 |
|  | 650 | 22 | 20 | 32* | 22 |
|  | 618 | 41.5 | 42 | 42 | 46 |
|  | 528 | 33 | 41* | 42* | 42* |
| *Prehension* | 242 | 47.5 | 45 | 49 | 49 |
| MDD = 6 | 192 | 42 | 47** | 44 | 46** |
| MCID = 3 | 650 | 12.5 | 12 | 17** | 16** |
|  | 618 | 15.5 | 18 | 21** | 19** |
|  | 528 | 42 | 42 | 48* ** | 44 |

**Supplementary Table 2.** Spinal Cord Independence Measure III (SCIM III) and Quality of Life (QLI-SCI) scores of each participant throughout the study. N.b. the baseline score was calculated as the average value between the scores of the two baseline sessions.

| Participant | 242 | 192 | 650 | 618 | 528 |
| --- | --- | --- | --- | --- | --- |
| *SCIM III* |  |  |  |  |  |
| Baseline | 51 | 64.5 | 53.5 | 25 | 83.5 |
| ULTP | 55 | 69 | 46 | 23 | 81 |
| ULTP+TCES | 55 | 67 | 58 | 25 | 90 |
| Follow-up | 49 | 67 | 54 | 26 | 81 |
| *QLI-SCI* |  |  |  |  |  |
| Baseline | 76.5 | 86.1 | 45.3 | 43.2 | 74.9 |
| ULTP | 79.9 | 86.6 | 54.9 | 47.2 | 82.5 |
| ULTP+TCES | 85.8 | 85.1 | 54.8 | 42.1 | 87.5 |
| Follow-up | 79.4 | 84.7 | 48.2 | 45.2 | 84.1 |

**Supplementary Table 3.** Fixed-effects table for the linear mixed model run on the force values produced during the grasping and lifting tasks. *η_p_*^2^ = partial eta squared.

| **Parameter** | **Numerator df** | **Denominator df** | ***F*** | **Sig.** | ***η*2** |
| --- | --- | --- | --- | --- | --- |
| Time | 3 | 6 | 7.868 | 0.017 | 0.80 |
| Order | 1 | 6 | 0.442 | 0.531 | 0.07 |
| Dominance | 1 | 6 | 0.137 | 0.724 | 0.02 |
| Time*Order | 3 | 6 | 8.796 | 0.013 | 0.81 |
| Time*Dominance | 3 | 6 | 6.108 | 0.030 | 0.75 |
| Dominance*Order | 1 | 6 | 3.719 | 0.102 | 0.38 |
| Time*Order*Dominance | 3 | 6 | 7.641 | 0.018 | 0.79 |
